# The Mask-Wearing Bias In The Estimates Of Vaccine Efficacy

**DOI:** 10.1101/2021.10.19.21265093

**Authors:** Andrew Matytsin

## Abstract

In the United States mask wearing is positively correlated with vaccine acceptance. This correlation introduces an important bias into real-world estimates of vaccine efficacy. I derive the formulae for vaccine efficacy that correct for this phenomenon and show that such biases explain some of the differences between higher estimates of vaccine efficacy observed in the US studies, on one hand, and lower estimates from Israel and Pfizer trials, on the other hand. Control for such biases is important for currently-debated public health decisions regarding COVID19 vaccine booster doses.

## 1 Overview

In the recent weeks, regulators and scientists have been vigorously debating the need for the third (“booster”) doses of COVID19 vaccines. At the center of the debate are the data from studies by Pfizer and collaborators [1, 2] and Israeli researchers [3, 6] showing that the efficacy of the Pfizer BNT162b2 COVID19 vaccine decreases after about 4-6 months. The need for boosters is questioned by other authors and regulators [7, 8, 9] who aver that the effectiveness of vaccines, especially against severe disease, appears to still be high in the United States [10, 11, 12]. Which view is correct has important consequences for the public health decisions currently being made all over the world.

In this note I argue that the discrepancy in estimated vaccine efficacy seen in various countries can be explained by a confounder not usually recognized as such^12^. Especially in the United States, there appears to be a strong correlation between mask wearing (and adherence to other infection-control measures)^3^ and vaccine acceptance [17]. Because the infection-control measures are highly useful in preventing COVID19, an observational US study that fails to control for mask wearing will yield an overly-optimistic estimate of vaccine effectiveness.

Among all countries, the United States stands out in how polarized the reactions to the threat of COVID19 are. At one extreme, one group, which we shall call “group R,” denies that COVID19 is a serious threat, refuses to wear masks and declines vaccines. The other group, “group D,” has a high vaccine acceptance rate and is more likely to follow infection control practices. Notably, there is a significant geographic clustering of persons in groups R and D that parallels to a degree the political preferences predominant in certain states.

It is clear that if the mask-vaccine correlation were perfect — that is, if there were only groups R and D, and the persons in R used neither masks nor vaccines, while the persons in D used both,—any observational estimate of vaccine efficacy would measure only the joint effectiveness of masking plus vaccination. Because masking is highly effective, any estimate from such studies would overestimate the efficacy of the vaccine alone. And because masks remain effective against more infectious variants (e.g., the Delta variant) the observational estimate would not decrease as fast, creating a false sense of safety.

In May of 2021 the US Centers for Disease Control and Prevention (CDC) issued guidance advising the fully-vaccinated that they did not need to wear masks, but that the unvaccinated did. Had this practice been followed universally, it would have introduced a negative correlation between mask wearing and vaccination status. That negative correlation would have led to a downward bias in real-world estimates of vaccine efficacy.

In other countries the degree to which the response to COVID19 is polarized appears to be less, and so we can expect the mask-vaccine correlation to be less pronounced. Moreover, the countries that enforce masking (especially for the unvaccinated), vaccine passports and other public health measures more effectively than the US can be expected to have zero or negative correlation between vaccination and avoidance of exposure^4^ to the SARS-Cov2 virus. For example, in a country that has a functional vaccination passport system the vaccinated would be able to attend large gatherings more than the unvaccinated. All of this would bias the real-wold estimates of vaccine effectiveness in such countries downward. It may well be that Israel is in that category.

In this paper we shall derive a very simple formula that shows by how much the real-world estimates need to be adjusted to control for the effect of masks. We shall see that the relative risk of a vaccinated person compared to the unvaccinated is to be multiplied by a factor that depends only on the effectiveness of masking. The adjustment factor does not depend on the vaccine effectiveness itself. Because the masks (together with other infection-avoidance techniques) are very effective, the adjustment factor is far from negligible and will affect significantly our estimates of risks to the vaccinated persons.

## 2 Corrected Estimate Of The Vaccine Effectiveness

In this section, we shall denote *RR* the relative risk of the vaccinated person developing the disease, compared to the unvaccinated person. Because the vaccinated are less likely to fall sick, *RR* < 1. We shall distinguish *RR*_obs_, the relative risk as measured from observational studies, from *RR*_true_, the true risk that, for example, could be determined from the properly controlled and blinded clinical trial. The more-frequently quoted *vaccine effectiveness* is then *V E* = 1 − *RR*.

We will show that

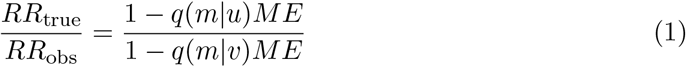

where *q*(*m*|*u*) is the proportion of those unvaccinated that choose to wear masks and *q*(*m*|*v*) is the proportion of mask-wearers among the vaccinated. *ME* is the effectiveness of masks that shows how much less often the mask-wearers develop the disease compared to those who do not wear masks.

Upon some reflection, this formula is obvious. If among the unvaccinated the fraction of *q*(*m*|*u*) wear masks, then *q*(*m*|*u*)*ME* is the fraction of the unvaccinated that would be spared the disease due to mask-wearing. Thus the factor *r*_*u*_ = 1 − *q*(*m*|*u*)*ME* shows the average risk among all unvaccinated relative to the risk of those unvaccinated and not-masked. Likewise, *r*_*v*_ = 1 − *q*(*m*|*v*)*ME* is the average risk of all vaccinated relative to those who are vaccinated but not masked. If *r*_*v*_ ≠ *r*_*u*_, the observed ratio of case rates between vaccinated and unvaccinated comes from two sources: the true effect of the vaccine on reducing the cases and the reduction due to the masking, which amounts to *r*_*v*_*/r*_*u*_. We give a formal derivation of (1) in the next section.

If masking (and other infection-control measures) were practiced equally by the vaccinated and not vaccinated, the adjustment would be unnecessary. Unfortunately, there is evidence that *q*(*m*|*u*) ≠ *q*(*m*|*v*). For example, a Gallup poll taken in June 2021 [13, 14] shows that in the United States, about 77% of the vaccinated continued to follow masking and other infection-control practices, whereas among the unvaccinated that percentage dropped to 38%. In our notation, *q*(*m*|*u*) = 0.38 and *q*(*m*|*v*) = 0.77. A detailed study of vaccine intentions [17]^5^ showed a similar or greater gap between those who intended to accept the vaccine and those who do not.

At any reasonable values of *ME* this leads to a significant adjustment. Using *ME* = 50% in (1) leads to an adjustment factor of about 1.4. That is, a naive estimate of the relative risk obtained from observational data would underestimate that risk by 40%. If *ME* is higher, the correction increases significantly. If *ME* = 0.8, the adjustment factor becomes 1.8, which means that the risk to the vaccinated is by about 80% greater than estimated. At *ME* = 90% the risk is 110% greater and in the extreme when masks are very effective (such as N95 masks) the adjustment factor can get to its upper limit of 2.7, almost *tripling* the estimated risk.

In Table 1 we present three examples of the bias adjustment for realistic parameter values.^6^ In each case, the adjustment is not negligible. But what matters more, is that while in the last two cases the naive estimate of vaccine efficacy is *conservative*, in the first case, relevant to the U.S. studies, the bias results in a potentially dangerous underestimate of risk.^7^ Unfortunately, the data sets used for observational studies do not contain much information about adherence to masking and other safety practices. I hope that the analysis in this paper will help to improve that situation. When possible — for example, in clinical trials and surveys — it would help to include questions about the participants’ behavior. In other observational studies, it would be desirable, at a minimum, to evaluate the dependence of estimates on various correlates of masking. These may include the counts of other respiratory infections, geographical location within the U.S. and, possibly, political orientation.^8^

**Table 1:** Examples of changes to vaccine effectiveness due to the masking bias.

| Scenario | Bias | $VE_{\text{obs}}$ | $RR_{\text{obs}}$ | $VE_{\text{true}}$ |
| --- | --- | --- | --- | --- |
| Observational study — US | Unvaccinated mask less than the vaccinated | 90% | 0.1 | 81% |
| | Assumptions: $q(m\vert u) = 0.4$, $q(m\vert v) = 0.8$, $ME = 0.8$ | 70% | 0.3 | 43% |
| Observational study — enforced mask mandate for unvaccinated, vaccine passports | Vaccinated may have more exposure to the virus | 90% | 0.1 | 95% |
| | Assumptions: $q(m\vert u) = 0.8$, $q(m\vert v) = 0.4$, $ME = 0.8$ | 70% | 0.3 | 84% |
| Randomized clinical trial with functional unblinding due to more side effects of the vaccine compared to placebo | Those receiving vaccine are less careful in avoiding exposure | 90% | 0.1 | 94% |
| | Assumptions: $q(m\vert u) = 0.9$, $q(m\vert v) = 0.7$, $ME = 0.8$ | 70% | 0.3 | 81% |

**Table 2:** The notation. Left table: the number of persons in each of the four categories obtained by classifying people as masked/unmasked and vaccinated/unvaccinated. Right table: the proportion of those who developed disease in these categories.

|  | unvaccinated | vaccinated |  | unvaccinated | vaccinated |
| --- | --- | --- | --- | --- | --- |
| no masks | $n_{\mathfrak{u}\mathfrak{u}}$ | $n_{\mathfrak{u}\mathfrak{v}}$ | no masks | $p_{\mathfrak{u}\mathfrak{u}}$ | $p_{\mathfrak{u}\mathfrak{v}}$ |
| masks | $n_{\mathfrak{m}\mathfrak{u}}$ | $n_{\mathfrak{m}\mathfrak{v}}$ | masks | $p_{\mathfrak{m}\mathfrak{u}}$ | $p_{\mathfrak{m}\mathfrak{v}}$ |

That *q*(*m*|*u*) *< q*(*m*|*v*) is a rather illogical situation that may distinguish the United States from other countries. On the contrary, it would be logical for the unvaccinated to keep wearing masks while for the vaccinated to drop some of the more burdensome precautions. That was the essence of the CDC guidance issued in May 2021 [18] and, had it been adhered to, we would be in the situation where *q*(*m*|*u*) *> q*(*m*|*v*). That would have the opposite effect on the bias in the estimates of vaccine effectiveness. Vaccines would appear *less* effective in observational studies than they in fact are.^9^ Countries which enforce vaccine passports, or otherwise limit to the vaccinated the admittance to certain venues, are also in this category. There, because the vaccinated have more opportunities to interact with others, they are subject to greater exposure to the virus. Israel is in this category. As a result, we can expect that observational studies from Israel do not overstate vaccine effectiveness and may, in fact, somewhat underestimate it.^10^

These results have significant implications for public health decisions debated now. Certain experts and regulators in the United States questioned the need for third vaccine doses, pointing, among other issues, to the higher estimates of vaccine effectiveness from observational studies performed with the US data, when compared to the data from Israel [7, 9]. The FDA briefing document on the third dose for the Prizer vaccine stated “Furthermore, US-based studies of post-authorization effectiveness of BNT162b2 may most accurately represent vaccine effectiveness in the US population.”[8] Our analysis suggests that it is particularly the US data that have to be used with great caution. Because of the strong mask-vaccination correlation, those relying on the US data alone may dangerously underestimate the true risk of breakthrough infections or, especially, hospitalizations.^11^ Some recently published data contain independent hints that this may be true. For example, the analysis in [19] shows higher estimated vaccine effectiveness among those *>* 65 years of age, compared to 18-49-olds. Even though that paper does not report the statistical significance of their estimates, the direction of the discrepancy is consistent with mask wearing being practiced more by the elderly, affecting the comparison between age groups.

## 3 Derivation Of The Adjustment Formula

In this section we derive the adjustment formula (1). We also define more precisely what is meant by vaccine efficacy when some wear masks but other do not and discuss a more general, and intriguing, possibility that the true vaccine efficacy itself may be different with and without masks. This section can be skipped by readers not concerned with mathematical details.

Imagine a population which is split by vaccination status and by mask-wearing practices. We then have four clases of persons, and we shall denote *n*_*mv*_ the number of persons in each class. The first index, *m* ∈ {`m, u`}, labels whether the person wears masks (`m` for “masked”) or not (u for “unmasked”) while the second index *v* ∈ {`v, u`}, shows whether the person is vaccinated (`v`) or not (`u`). Likewise, we shall denote *p*_*mv*_ the proportion of those developing disease in each of these categories.

With this notation, we can define *vaccine efficacy among those using masks*,

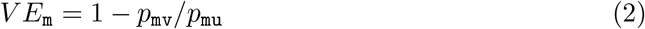

and the *vaccine efficacy among those not using masks*,

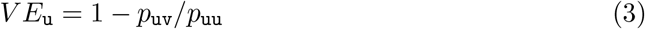

Obviously, if the vaccine efficacy does not depend on whether one wears a mask, then *V E*_m_ = *V E*_u_ ≡ *V E*. But it is possible that these two vaccine effectiveness measures are different. For example, if a vaccine is able to counteract only a moderate amount of the viral challenge but then fails if one is exposed to a greater amount of the virus, then we may expect *V E*_m_ *> V E*_u_. I am not aware of an empirical analysis that addresses that question for SARS-Cov2, but it would be interesting to perform such.

It is easy to derive the adjustment formula for the more general case anyway. Among the vaccinated, the total number of cases is *n*_uv_*p*_uv_ + *n*_mv_*p*_mv_. The rate of disease per-person among the vaccinated is therefore

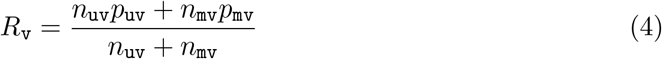

Among unvaccinated, the rate of disease per person is, similarly,

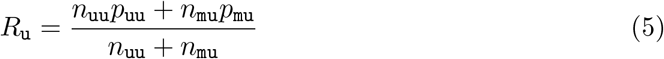

The naively measured relative risk is then

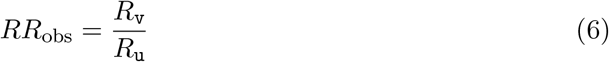

with the vaccine efficacy being, by definition,

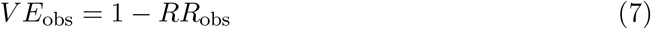

The true vaccine efficacy for the population where some use masks and some do not is, however, different. It must be defined very carefully. Imagine a clinical trial where both the vaccine arm and the control arm have *n*_uv_ participants that do not wear masks and *n*_mv_ participants who do. That is, in both arms the pattern of mask wearing is the same as the actual pattern in the vaccinated population.^12^ Then the number of cases in the vaccine arm would be expected to be *n*_uv_*p*_uv_ + *n*_mv_*p*_mv_. In the control arm we would expect different proportions of disease, with the resulting case count *n*_uv_*p*_uu_ + *n*_mv_*p*_mu_. The true relative risk for the vaccinated is then

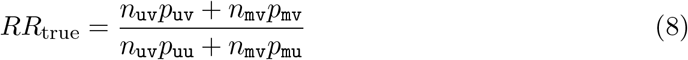

We can now evaluate the adjustment factor necessary to compute the true risk from the observed risk. Using (4,5,6) and (8),

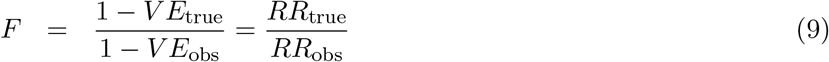

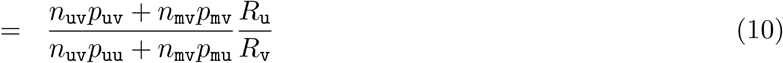

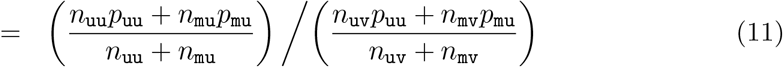

The respective proportions of mask-wearers among the vaccinated and unvaccinated are

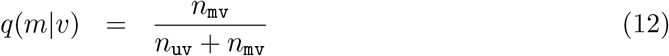

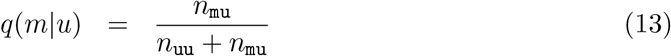

Using these definitions and (11),

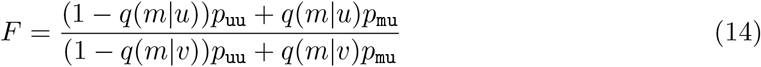

Note that this expression depends only on the ratio *p*_mu_*/p*_uu_, which is directly related to the *effectiveness of masks* (absent vaccination). In fact, similarly to *V E*, we can define the mask effectiveness

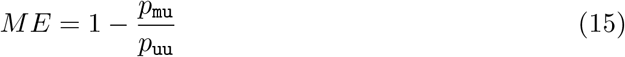

In terms of *ME*,

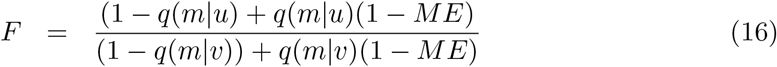

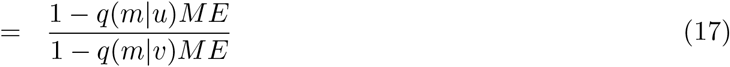

which is our desired result (1).

It pays to analyze the expression for the true vaccine efficacy more closely. From (8)

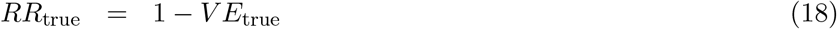

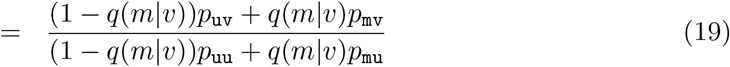

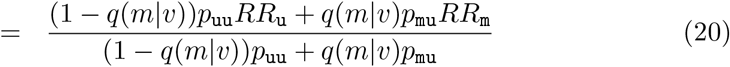

where, consistent with (2) and (3), we use the relative risks of disease for vaccinated with and without masks

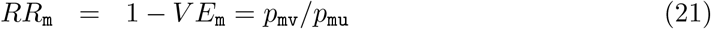

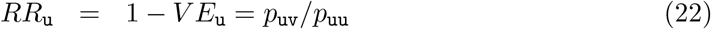

Given (15), *p*_mu_ = *p*_uu_(1−*ME*) and we see that the true relative risk of disease is a weighted average of *RR*_u_ and *RR*_m_,

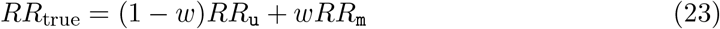

with the weight

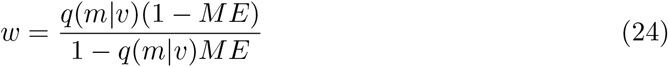

Because 0 ≤ *q*(*m*|*v*) ≤ 1 and 0 ≤ *ME* ≤ 1, it is always true that 0 ≤ *w* ≤ 1.

Obviously, if the vaccine efficacy is the same with and without masks, then *RR*_true_ = *RR*_m_ = *RR*_u_ and the true vaccine efficacy does not depend on *w*. Also, if *ME* = 1 (masks are perfectly effective) then *w* = 0 and *RR*_true_ = *RR*_u_. That is also correct: in the extreme case when masks filter out all of the virus those masked never develop the disease and thus do not contribute to the case counts. Hence, the measurement of vaccine efficacy comes from only those unmasked. Finally, if *ME* = 0, *w* = *q*(*m*|*v*) and the relative risk is simply the weighted average of the risks with and without masks, with the weight equal to the proportion of those wearing masks.

One very interesting feature of (24) is that *w < q*(*m*|*v*) (except for the trivial cases when *q*(*m*|*v*) is either 0 or 1, or *ME* = 0.) The reason for this is that because the masked have extra protection, the case counts they are contributing are relatively smaller than the case counts coming from the unmasked. Thus, the weight of the vaccine efficacy coming from the masked is depressed compared to their proportion in the population.

As we have already mentioned, it would be very interesting to see whether *V E*_u_ *< V E*_m_. That could be so if the vaccines are better able to neutralize a smaller quantity of the virus (inhaled through the mask) than a larger quantity, to which a person without a mask would be exposed. This effect would be an additional source of interaction between masking and vaccination. The formula for bias in (1) is robust to such details and is valid whether or not *V E*_u_ = *V E*_m_.

## 4 Conclusions

Because adherence to masking and acceptance of vaccines are correlated, at least in the United States, vaccine efficacy estimated from real-world data may be overstated by a significant amount. The greater the correlation or the mask effectiveness, the more the bias. We have derived the adjustment formulas that recover the true vaccine efficacy if the proportions of mask wearers are known. We showed that, for realistic parameters, the masking bias could lead to a significant underestimate of the risk of disease. Therefore, it is imperative that future studies of vaccines seek to quantify or at least control for differential rates of mask wearing between the vaccinated and the unvaccinated.

These conclusions have implications for the current debate on booster doses. The skepticism about the need for the third dose rests largely on the existence of studies that show continued protection against severe disease from just two doses. Our analysis shows that this view may understate the risk. It also suggests that using data from another country that is not so polarized may provide better guidance as to the true vaccine effectiveness.

## Data Availability

All data produced in the present work are contained in the manuscript

## Footnotes

1 Recognized reasons for differences include the use of Moderna vaccine in the US, different definitions of severe disease and different speed of the vaccine rollout. That said, [12] mentions the lack of control for mask use as a general limitation of the study, without discussing its potential effect on the results. To my knowledge, the idea that the mask-vaccine correlation leads to a systematic bias, rather than a random error, has not been studied. Certainly, it has not been discussed in the advisory committee proceedings on the Pfizer vaccine boosters.

2 For example, a comprehensive presentation [5] at the FDA hearing on the booster doses was devoted entirely to the question of bias in real-world studies, but made no mention of this effect.

3 Thought this paper, when we speak of mask wearing, we include all other infection-avoidance techniques, such as physical distancing, avoidance of crowded conditions or indoor spaces, isolation and the like.

4 Thus, a positive correlation between the vaccination and exposure *level*.

5 That study was done in 2020, before the vaccines were available. However, its conclusions agree well with the observation that vaccine rejectors are about half as likely to wear a mask. For other infection-control practices, that study reports an even greater gap (Table 1 of [17].)

6 We used *ME* = 0.8 because the evidence review in [16] points to risk ratios around 0.2 for a variety of infection-avoidance strategies, such as masking, physical distancing and eye protection. An even larger *ME* is suggested by the data in that reference for N95 masks. Of course, depending on the type of masks used and the level of adoption in the community the final *ME* may vary greatly (see also [15].) The main assertion of this paper is that for any positive *ME* there will be a *systematic bias* in *V E* estimates, rather than a random error. For example, if we use *ME* = 0.5 instead of 0.8 in Table 1, the observed *V E* = 70% from a US observational study would be adjusted down to 60%, still an appreciable amount.

7 With respect to clinical trials, the masking bias was discussed in general press [20].

8 It is documented that both vaccine hesitancy and mask resistance in the U.S. are correlated with political orientation [17]. I am not aware of studies documenting a similarly large split elsewhere.

9 It is ironic that by rejecting masks the anti-vaccine crowd has succeeded in biasing the estimates of vaccine effectiveness *upward* and thus leading some in the CDC to believe that the third doses are unnecessary for the general population.

10 A very recent study from Israel [4] has a detailed analysis of behavior around the time of (third) vaccination, addressing the biases described in [5]. Because that study compares the persons who already had two doses to those who got three, it is unlikely to be affected by ardent vaccine and mask rejectors.

11 Our methodology applies with equal force to any definition of disease, be it positive tests, symptomatic infection, hospitalization, or death. The only difference is that the proper value of *ME* should be used, which may be different for different disease definitions. Notably, for severe disease, *ME* can be expected to be larger [22, 23], leading to a larger bias.

12 This last point is essential. The control arm must have the same proportion of mask wearers, because otherwise we are not controlling properly for mask use.

## References

[1] Prizer Inc., Vaccines and Related Biological Products Advisory Committee September 17, 2021 Meeting Briefing Document—Sponsor, https://www.fda.gov/media/152161/download.

[2] Sara Y Tartof, Jeff M Slezak, Heidi Fischer, et al. Effectiveness of mRNA BNT162b2 COVID-19 vaccine up to 6 months in a large integrated health system in the USA: a retrospective cohort study. Lancet, Published Online October 4, 2021, https://doi.org/10.1016/S0140-6736(21)02183-8.

[3] Haas EJ, Angulo FJ, McLaughlin JM, et al. Impact and effectiveness of mRNA BNT162b2 vaccine against SARS-CoV-2 infections and COVID-19 cases, hospitali-sations, and deaths following a nationwide vaccination campaign in Israel: an observational study using national surveillance data. Lancet 2021; 397: 1819–29.

[4] Yinon M. Bar-On, Yair Goldberg et al., BNT162b2 vaccine booster dose protection: A nationwide study from Israel, https://doi.org/10.1101/2021.08.27.21262679

[5] Jonathan Sterne, Vaccines and Related Biological Products Advisory Committee September 17, 2021 Meeting Presentation — Real-world effectiveness of COVID-19 Vaccines, https://www.fda.gov/media/152241/download.

[6] Sharon Alroy-Preis and Ron Milo, Vaccines and Related Biological Products Advisory Committee September 17, 2021 Meeting Presentation — Booster Protection against confirmed infections and severe disease — data from Israel, https://www.fda.gov/media/152205/download.

[7] Philip R Krause, Thomas R Fleming, Richard Peto, et al. Considerations in boosting COVID-19 vaccine immune responses. Lancet, Published Online September 13, 2021 https://doi.org/10.1016/S0140-6736(21)02046-8.

[8] U.S. Food and Drug Administration, Vaccines and Related Biological Products Advisory Committee September 17, 2021 Meeting Briefing Document — FDA, https://www.fda.gov/media/152176/download, pages 8–9.

[9] Yasmeen Abutaleb and Lena H. Sun, How CDC data problems put the U.S. behind on the delta variant. Critics say the CDC’s failure to share real-time data led to overly rosy assessments of vaccine effectiveness — and complacency on the part of many Americans, Washington Post, August 19, 2021, https://www.washingtonpost.com/health/2021/08/18/cdc-data-delay-delta-variant/

[10] Scobie HM, Johnson AG, Suthar AB, et al. Monitoring Incidence of COVID-19 Cases, Hospitalizations, and Deaths, by Vaccination Status — 13 U.S. Jurisdictions, April 4–July 17, 2021. MMWR Morb Mortal Wkly Rep 2021;70:1284–1290. DOI: http://dx.doi.org/10.15585/mmwr.mm7037e1

[11] Grannis SJ, Rowley EA, Ong TC, et al. Interim Estimates of COVID-19 Vaccine Effectiveness Against COVID-19–Associated Emergency Department or Urgent Care Clinic Encounters and Hospitalizations Among Adults During SARS-CoV-2 B.1.617.2 (Delta) Variant Predominance — Nine States, June–August 2021. MMWR Morb Mortal Wkly Rep 2021;70:1291–1293. DOI: http://dx.doi.org/10.15585/mmwr.mm7037e2.

[12] Bajema KL, Dahl RM, Prill MM, et al. Effectiveness of COVID-19 mRNA Vaccines Against COVID-19–Associated Hospitalization — Five Veterans Affairs Medical Centers, United States, February 1–August 6, 2021. MMWR Morb Mortal Wkly Rep 2021;70:1294–1299. DOI: http://dx.doi.org/10.15585/mmwr.mm7037e3

[13] Lydia Saad, Strict Social Distancing in the U.S. Dwindles to 18%, July 8, 2021, https://news.gallup.com/poll/352166/strict-social-distancing-dwindles.aspx

[14] Alison Durkee, Mask Wearing Plummets — Especially Among Unvaccinated - But Delta Variant Creates Growing Anxiety About Covid-19, Polls Find. Forbes, July 8, 2021.

[15] Howard J, Huang A, Li Z et al., An evidence review of face masks against COVID-19, PNAS January 26, 2021 118 (4) e2014564118; https://doi.org/10.1073/pnas.2014564118

[16] Derek K Chu, Elie A Akl et al. on behalf of the COVID-19 Systematic Urgent Review Group Effort (SURGE) study authors, Physical distancing, face masks, and eye protection to prevent person-to-person transmission of SARS-CoV-2 and COVID-19: a systematic review and meta-analysis, Lancet 2020; 395: 1973–87, PublishedOnline June 1, 2020 https://doi.org/10.1016/S0140-6736(20)31142-9

[17] Latkin CA, Dayton L, Yi G, Colon B, Kong X (2021) Mask usage, social distancing, racial, and gender correlates of COVID-19 vaccine intentions among adults in the US. PLoS ONE 16(2): e0246970. https://doi.org/10.1371/journal.pone.0246970

[18] Your Guide to Masks, Centers for Disease Control and Prevention, https://www.cdc.gov/coronavirus/2019-ncov/prevent-getting-sick/about-face-coverings.html. That guidance has been updated since.

[19] Rosenberg ES, Holtgrave DR, Dorabawila V, et al. New COVID-19 Cases and Hospitalizations Among Adults, by Vaccination Status — New York, May 3–July 25, 2021. MMWR Morb Mortal Wkly Rep 2021;70:1306–1311. DOI: http://dx.doi.org/10.15585/mmwr.mm7037a7, Figure 1.

[20] Aria Bendix and Lauren Lee, Vaccine trials didn’t monitor one variable: volunteers’ behavior. ‘Masks and social distancing were left up to us,’ a participant said. Business Insider, November 25, 2020, https://www.businessinsider.com/vaccine-trials-pfizer-moderna-didnt-regulate-participants-masks-social-distancing-2020-11

[21] Benjamin Rader, Laura F White et al., Mask-wearing and control of SARS-CoV-2 transmission in the USA: a cross-sectional study Lancet Digit Health 2021; 3: e148–57 Published Online January 19, 2021 https://doi.org/10.1016/S2589-7500(20)30293-4

[22] Monica Gandhi, Chris Beyrer and Eric Goosby, Masks Do More Than Protect Others During COVID-19: Reducing the Inoculum of SARS-CoV-2 to Protect the Wearer. J Gen Intern Med 35(10):3063–6, DOI: 10.1007/s11606-020-06067-8

[23] Monica Gandhi and George W. Rutherford, Facial Masking for Covid-19 — Potential for “Variolation” as We Await a Vaccine, N Engl J Med 383;18, http://nejm.org, October 29, 2020

